# The analysis of the long-term impact of SARS-CoV-2 on the cellular immune system in individuals recovering from COVID-19 reveals a profound NKT cell impairment

**DOI:** 10.1101/2020.08.21.20179358

**Authors:** Jia Liu, Xuecheng Yang, Hua Wang, Ziwei Li, Hui Deng, Jing Liu, Shue Xiong, Junyi He, Chunxia Guo, Weixian Wang, Gennadiy Zelinskyy, Mirko Trilling, Ulf Dittmer, Mengji Lu, Kathrin Sutter, Tina Senff, Christopher Menne, Joerg Timm, Yanfang Zhang, Fei Deng, Xuemei Feng, Yinping Lu, Jun Wu, Dongliang Yang, Baoju Wang, Xin Zheng

## Abstract

The *coronavirus disease 2019* (COVID-19) pandemic caused by the *severe acute respiratory syndrome coronavirus 2* (SARS-CoV-2) affects millions of people and killed hundred-thousands of individuals. While acute and intermediate interactions between SARS-CoV-2 and the immune system have been studied extensively, long-term impacts on the cellular immune system remained to be analyzed. Here, we comprehensively characterized immunological changes in peripheral blood mononuclear cells in 49 COVID-19 convalescent individuals (CI) in comparison to 27 matched SARS-CoV-2 unexposed individuals (UI). Despite recovery from the disease for more than 2 months, CI showed significant decreases in frequencies of invariant NKT and NKT-like cells compared to UI. Concomitant with the decrease in NKT-like cells, an increase in the percentage of Annexin V and 7-AAD double positive NKT-like cells was detected, suggesting that the reduction in NKT-like cells results from cell death months after recovery. Significant increases in regulatory T cell frequencies, TIM-3 expression on CD4 and CD8 T cells, as well as PD-L1 expression on B cells were also observed in CI, while the cytotoxic potential of T cells and NKT-like cells, defined by GzmB expression, was significantly diminished. However, both CD4 and CD8 T cells of CI showed increased Ki67 expression and were fully capable to proliferate and produce effector cytokines upon TCR stimulation. Collectively, we provide the first comprehensive characterization of immune signatures in patients recovering from SARS-CoV-2 infection, suggesting that the cellular immune system of COVID-19 patients is still under a sustained influence even months after the recovery from disease.

## Introduction

The sudden emergence and rapid global spread of *severe acute respiratory syndrome coronavirus 2* (SARS-CoV-2) and the resulting *coronavirus disease 2019* (COVID-19) poses an unprecedented health crisis to humankind. As of August 14^th^, 2020, there were more than 20 million documented cases of SARS-CoV-2 infection and more than 750 thousand individuals lost their lives. SARS-CoV-2-infected people exhibit a wide spectrum of disease manifestations ranging from moderate or even unnoticed symptoms (1) to life-threatening acute infections predominantly affecting the respiratory tract (2) but also other organs such as the kidney (3) and the central nervous system (4) can be harmed. Moderate cases show symptoms of fever, dry cough, fatigue, abnormal chest CT findings but with a good prognosis (5, 6). Conversely, some patients suddenly deteriorate towards acute respiratory distress syndrome (ARDS) or multiple organ failure, with fatality rates approaching 60% (7).

Recent studies demonstrated that SARS-CoV-2 infections strongly shape the immune system and result in its dysregulation, including imbalanced antiviral and pro-inflammatory responses, altered numbers and impaired functions of different immune cell subsets (8). We and others previously showed that lymphopenia and an inflammatory cytokine storm can be observed in COVID-19 patients and that their extents correlate with COVID-19-associated disease severity and mortality (9–12). The recovery of T cell counts and the end of the inflammatory cytokine storm in severe COVID-19 cases have been associated with a favorable disease outcome (9). However, the impact of SARS-CoV-2 on the cellular immune system after the recovery from the disease remains largely unknown.

Applying multi-color flow cytometry, we comprehensively characterized immunological changes in peripheral blood mononuclear cells (PBMCs) in 49 convalescent SARS-CoV-2-infected individuals (CI) in comparison to 27 matched SARS-CoV-2-unexposed individuals (UI). To our knowledge, our results provide the first in-depth description of immune signatures in the aftermath of SARS-CoV-2 infections in convalescent patients. Our data suggest that the immune system remains heavily influenced months after resolving SARS-CoV-2 infection.

## Methods

### Subjects

Thirty convalescent individuals who resolved their SARS-CoV-2 infection and a matched group comprising 21 SARS-CoV-2-unexposed individuals were recruited at the Department of Infectious Diseases, Union Hospital, Tongji Medical College, Huazhong University of Science and Technology and the Department of Gastroenterology from May to June 2020. The diagnosis of COVID-19 was based on the Guidelines for Diagnosis and Treatment of Corona Virus Disease 2019 issued by the National Health Commission of China (7^th^ edition, http://www.chinacdc.cn/jkzt/crb/zl/szkb_11803/jszl_11815/202003/t20200305_214142.html. Informed written consent was obtained from each patient and the study protocol was approved by the local medical ethics committee of Union Hospital, Tongji Medical College, Huazhong University of Science and Technology in accordance with the guidelines of the Declaration of Helsinki (2020IEC-J-587). Invariant NKT cell analysis was performed in a German cohort which has 19 CI and 6 UI. Written informed consent was given from each included individual and the study was approved by the ethics committee of medical faculty of the Heinrich Heine University Düsseldorf Germany (study number: 5350).

### Preparation of PBMCs

Peripheral blood mononuclear cells (PBMCs) of SARS-CoV-2-unexposed individuals and patients were isolated using Ficoll density gradient centrifugation (DAKEWE Biotech, Beijing) and were rapidly assessed by flow cytometry analysis without intermittent cryo-conservation.

### Flow cytometry

Surface and intracellular staining for flow cytometry analysis were performed as described previously(13, 14). For surface staining, cells were incubated with relevant fluorochrome-labeled antibodies for 30 min at 4°C in the dark. For intracellular cytokine staining, cells were fixed and permeabilized using the Intracellular Fixation & Permeabilization Buffer Set (Invitrogen, USA) and stained with APC-anti-IFN-γ, PerCP-Cy5.5-anti-IL-2, or FITC-anti-TNFα (BD Biosciences, USA). Freshly isolated cells were used for all assays. Approximately 100,000 PBMCs were acquired for each sample using a BD FACS Canto II flow cytometer. Data analysis was performed using FlowJo software V10.0.7 (Tree Star, Ashland, OR, USA). Cell debris and dead cells were excluded from the analysis based on scatter signals and Fixable Viability Dye eFluor 506.

### Analysis of effector T cell responses

PBMCs were resuspended in complete medium (RPMI 1640 containing 10% fetal calf serum, 10U/ml penicillin, 100μg/ml streptomycin, and 100μM 4-[2-hydroxyethyl]-1-piperazine ethanesulfonic acid buffer) and stimulated with by anti-CD3 (1μg/ml; BD Biosciences, USA), anti-CD28 (1μg/ml; BD Biosciences, USA), and recombinant interleukin-2 (20U/ml; Hoffmann-La Roche, Italy). Fresh medium containing IL-2 was added every 72 hours. On day 5, brefeldin A (BD Biosciences, San Diego, CA) was added to the media for 6 hours. Cells were washed and tested for Ki67 expression and secretion of IFN-γ, IL-2, and TNF-α by intracellular cytokine staining and subsequent flow cytometry analyses.

### Statistical Analysis

Statistics comparing two groups were done using the Mann-Whitney t-test. When more than two groups were compared, a one-way ANOVA was used with a Tukey post-hoc test (GraphPad Prism software; GraphPad Software Inc., San Diego, USA).

## Results

### Characteristics of the study cohort

To characterize the cellular immune system in individuals that had recovered from COVID-19, blood samples were analyzed about 3.5 months (Chinese cohort) or 1.5 months (German cohort) after the first diagnosis. The demographic profiles of the Chinese cohort are shown in Table 1. All CI had been diagnosed as either moderate(83.3%, 25/30) or mild COVID-19 cases (16.7%, 5/30). The median period between the first diagnosis of COVID-19 and blood sampling was 112 days (range: 60 to 136 days). Among all COVID-19 cases, 43.3% (13/30) were hospitalized and 23.3% (7/30) received oxygen inhalation treatment. Leukopenia and lymphopenia were observed in 43.5% (10/23) and 60.9% (14/23) of tested cases, respectively. Increased C-reactive protein and IL-6 levels were observed in 52.6% (10/19) and 76.9% (10/13) of tested patients, respectively. All moderate cases showed abnormal radiological findings suggesting pneumonia by chest computed tomography (CT) scans, while mild cases showed no radiological abnormality in the lungs. Twelve moderate cases and one mild case (43.3%, 13/30) had positive RT-PCR results for viral RNA. All patients were anti-SARS-CoV-2 IgM and/or IgG seropositive. At the time of blood sampling, 20% (6/30) of cases exhibited virus-specific IgM and IgG, 70% (21/30) were IgG single positive, and 10% (3/30) were IgM and IgG negative. The demographic profiles of the German cohort are shown in Table 2. This cohort has 19 CI, in which 10.53 % (2/19) were hospitalized. Nine cases (47.37%, 9/19) had positive RT-PCR results for viral RNA, and 18 cases (94.74%, 18/19) were anti-SARS-CoV-2 IgA and/or IgG seropositive. The median period between the first diagnosis of COVID-19 and blood sampling in the German cohort was 41 days. Only the analysis of invariant NKT (iNKT) cells was performed in the German cohort, while analysis of all other cell populations was performed in the Chinese cohort.

**Table 1.**
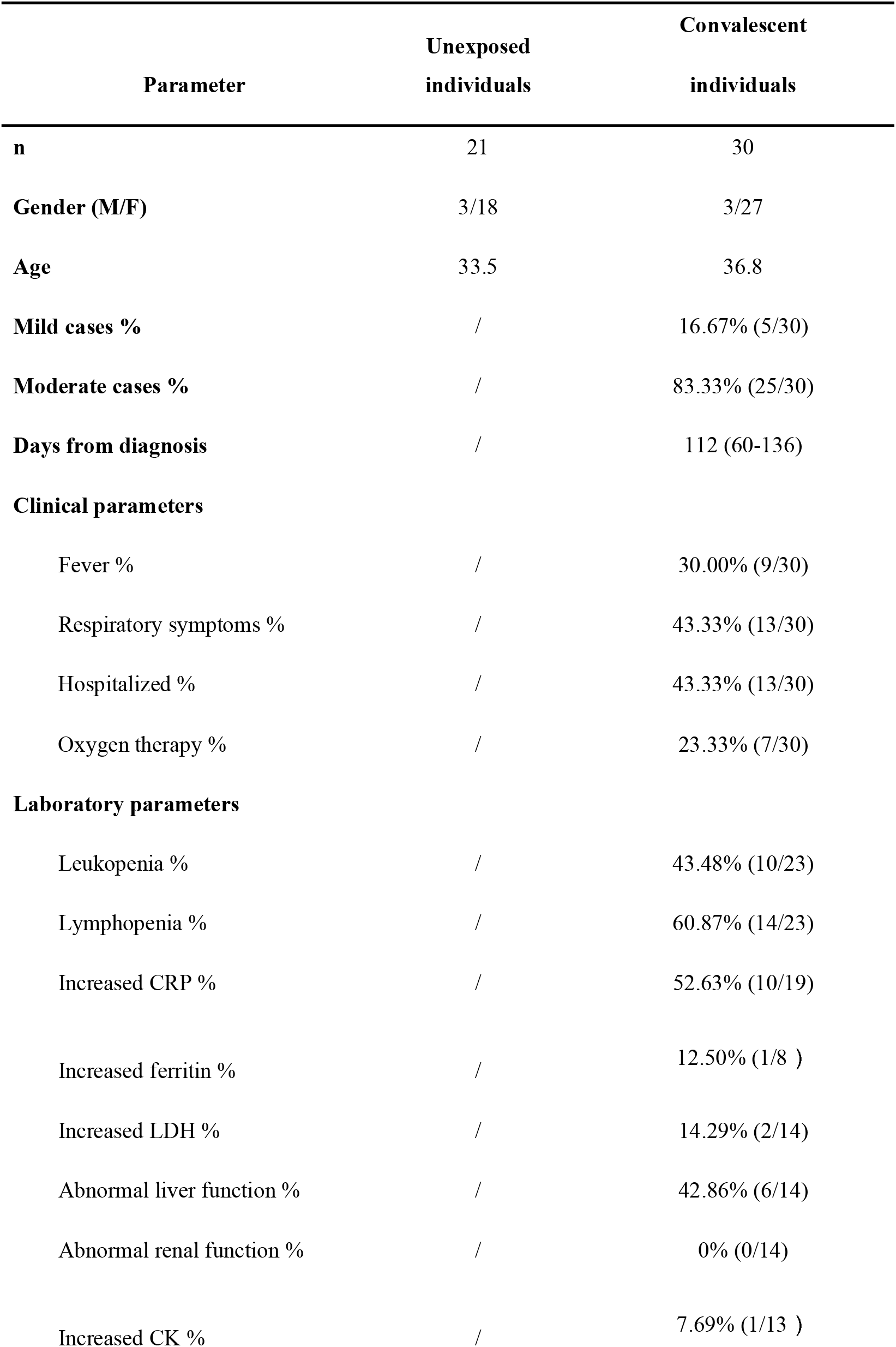
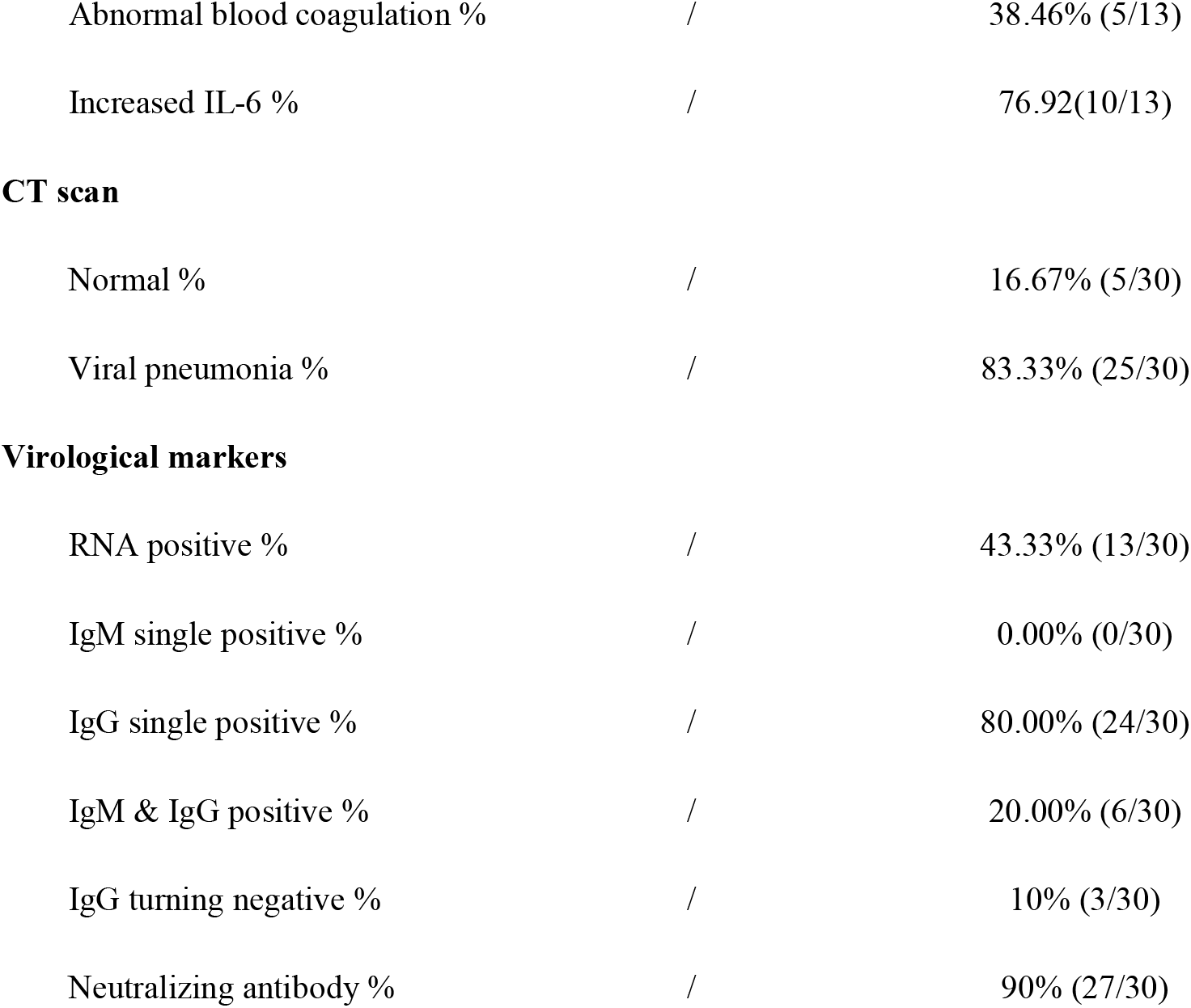
Baseline characteristics of the Chinese cohort

| Parameter | Unexposed<br>individuals | Convalescent<br>individuals |
| --- | --- | --- |
| **n** | 21 | 30 |
| **Gender (M/F)** | 3/18 | 3/27 |
| **Age** | 33.5 | 36.8 |
| **Mild cases %** | / | 16.67% (5/30) |
| **Moderate cases %** | / | 83.33% (25/30) |
| **Days from diagnosis** | / | 112 (60-136) |
| **Clinical parameters** |  |  |
| Fever % | / | 30.00% (9/30) |
| Respiratory symptoms % | / | 43.33% (13/30) |
| Hospitalized % | / | 43.33% (13/30) |
| Oxygen therapy % | / | 23.33% (7/30) |
| **Laboratory parameters** |  |  |
| Leukopenia % | / | 43.48% (10/23) |
| Lymphopenia % | / | 60.87% (14/23) |
| Increased CRP % | / | 52.63% (10/19) |
| Increased ferritin % | / | 12.50% (1/8 ) |
| Increased LDH % | / | 14.29% (2/14) |
| Abnormal liver function % | / | 42.86% (6/14) |
| Abnormal renal function % | / | 0% (0/14) |
| Increased CK % | / | 7.69% (1/13 ) |
| Abnormal blood coagulation % | / | 38.46% (5/13) |
| Increased IL-6 % | / | 76.92(10/13) |
| **CT scan** |  |  |
| Normal % | / | 16.67% (5/30) |
| Viral pneumonia % | / | 83.33% (25/30) |
| **Virological markers** |  |  |
| RNA positive % | / | 43.33% (13/30) |
| IgM single positive % | / | 0.00% (0/30) |
| IgG single positive % | / | 80.00% (24/30) |
| IgM & IgG positive % | / | 20.00% (6/30) |
| IgG turning negative % | / | 10% (3/30) |
| Neutralizing antibody % | / | 90% (27/30) |

**Table 2.** Baseline characteristics of the German cohort

| **Parameter** | **Unexposed**<br>**individuals** | **Convalescent**<br>**individuals** |
| --- | --- | --- |
| **n** | 6 | 19 |
| **gender (M/F)** | 2/4 | 10/9 |
| **age (mean)** | 46.3 | 44.8 |
| **Hospitalized %** |  | 10.53% (2/19) |
| **RNA positive** | / | 47.37% (9/19) |
| **Anti-SARS-CoV-2 IgA and/or IgG**<br>**positive\*** | / | 94.74% (18/19) |

\*OD ratio $\geq 1.1$ for IgA and IgG was considered positive (EUROIMMUN ELISA); index (S/C) $\geq 1.4$ is considered positive (Abbott CMIA)

The criteria for COVID-19 convalescent are as follows: afebrile for more than 3 days, resolution of respiratory symptoms, substantial improvement of chest CT images and two consecutive negative RT–qPCR tests for viral RNA in respiratory tract swab samples obtained at least 24 h apart. At time of blood sampling, all CI were negative for viral RNA test and had no recognized medical conditions.

### Characterization of immune cell subsets in individuals recovering from COVID-19

First, we characterized whether the overall immune cell composition in PBMCs differ between CI and UI by flow cytometry (as depicted in figure S1A and S1B). We observed that the profile of the immune cell composition of CI was distinct from that of UI (Figure 1A). Specifically, the frequencies but not the absolute numbers of CD4+ T cells in CI were slightly but significantly higher than in UI (Figure 1B and S1C), while no significant differences in absolute numbers and frequencies of total T cells, CD8+ T cells, B cells, NK cells, and monocytes were observed between CI and UI (Figure 1B and S1C). Interestingly, CI showed dramatic decreases in frequencies of both the NKT-like cell population (CD3+ CD56+) and the iNKT cell population (α-GalCer-CD1d tetramer+ and TCR Vα24-Vβ11+) compared to UI (Figure 1C and 1D). The absolute numbers of NKT-like cells of CI (median: 20.7/μ1) were only about 60% of the level observed in UI (median: 34.5/μ figure 1C). Besides, CI showed a significant increase in dendritic cells (DCs) in both the absolute numbers and frequencies compared to UI (Figure 1E).

**Figure 1.**
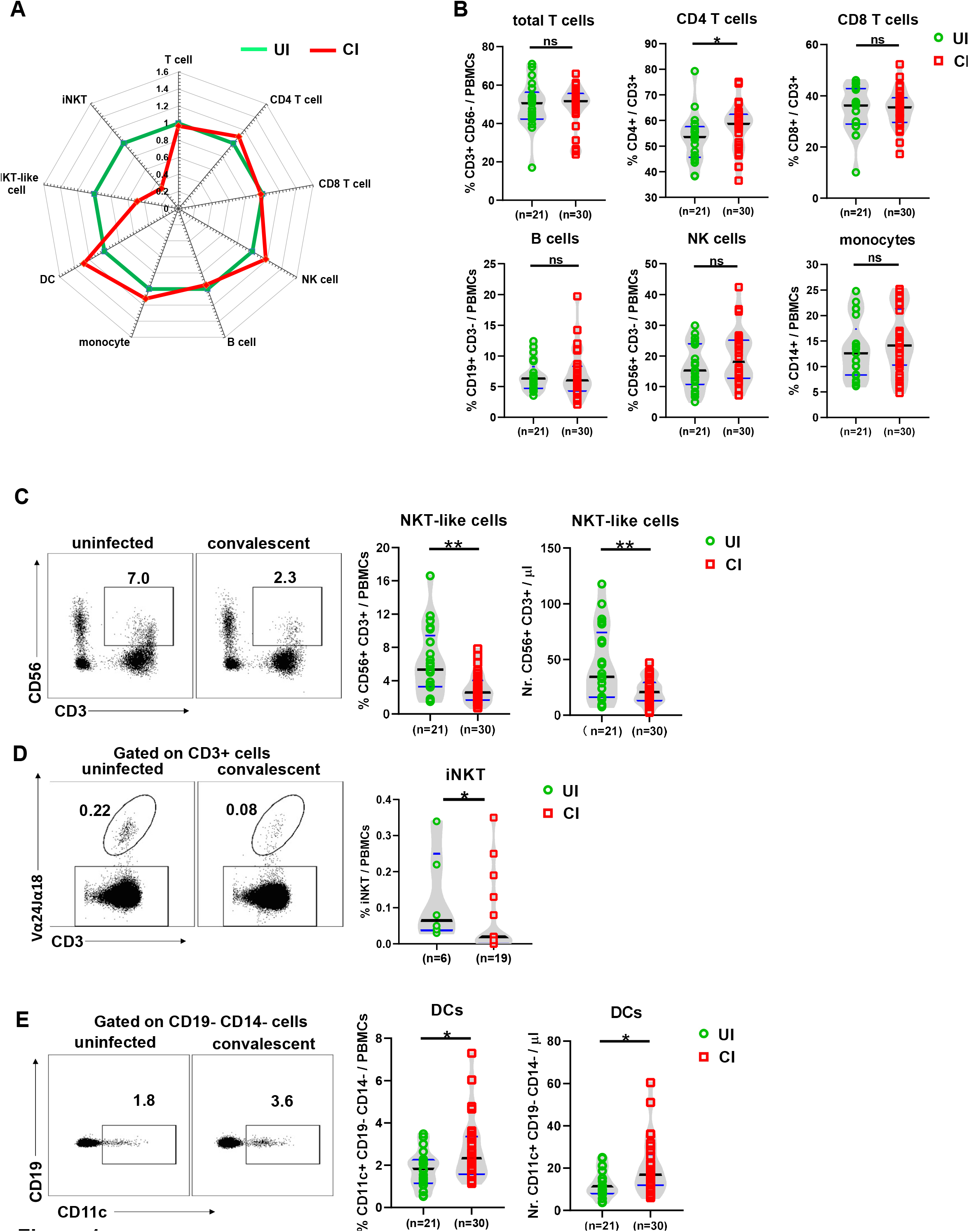
Characterization of immune cell subsets in individuals recovering from COVID-19. (A) The fold changes of the percentages (median) of total T cells, CD4 and CD8 T cells, B cells, NK cells, NKT-like cells, dendritic cells, and monocytes in the blood of CI (n=30) compared to those of UI (n=21) are depicted by radar plots. (B) The percentages of total T cells, CD4 and CD8 T cells, B cells, NK cells, and monocytes in the blood of UI (n=21) and CI (n=30) were analyzed by flow cytometry. C) The absolute numbers and percentages of NKT-like cells in the blood of UI (n=21) and CI (n=30) were analyzed by flow cytometry. (D) The percentages of invariant NKT (iNKT) cells in the blood of UI (n=6) and CI (n=19) were analyzed by flow cytometry. (E) The absolute numbers and percentages of dendritic cells in the blood of UI (n=21) and CI (n=30) were analyzed by flow cytometry.CI: COVID-19 convalescent individuals, UI: SARS-CoV-2-unexposed individuals. Black and blue lines in the figures: median and quartiles. Statistically significant differences are indicated by asterisks (* < 0.05, ** < 0.01; Non-parametric Mann-Whitney test).

Next, we examined whether the decrease of NKT-like cells in CI was associated with increased cell death. Annexin V and 7-AAD stainings were performed to analyze apoptosis and necroptosis of NKT-like cells, CD4 and CD8 T cells, B cells, and NK cells. A profound and significant increase in the frequency of the Annexin V and 7-AAD double positive NKT-like cells was observed in CI (median: 3.3%) compared to UI (median: 1.2%) (Figure 2A), suggesting that increased proportions of NKT-like cells of CI are undergoing apoptosis and/or necroptosis even after recovery from COVID-19. Importantly, the intensities of NKT-like cell death were inversely correlated with NKT cell frequencies in CI (Figure 2B). Besides, apoptosis and/or necroptosis of CD4+ T cells and B cells were also slightly increased in CI compared to those in UI (Figure 2C).

**Figure 2.**
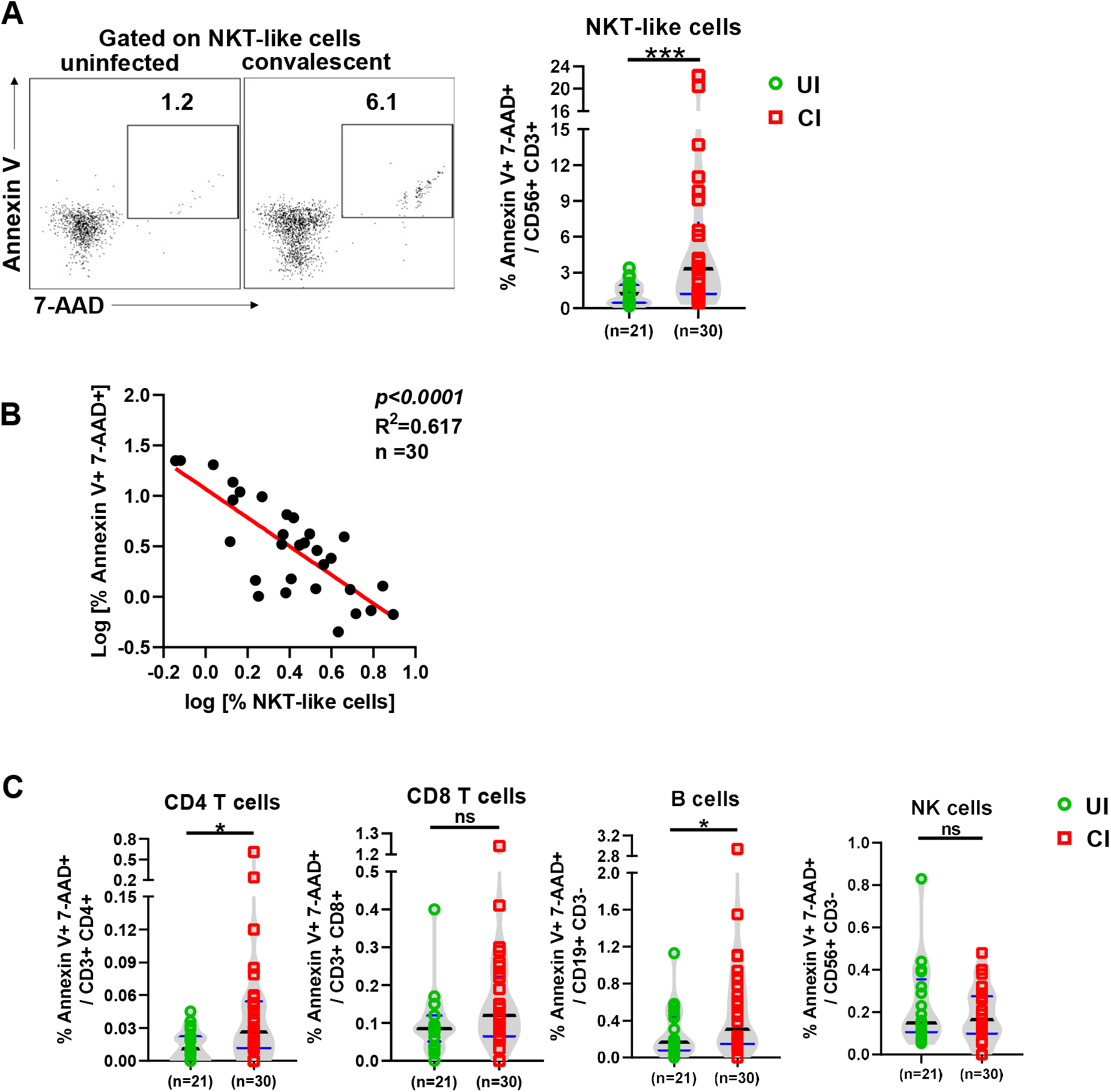
Characterization of immune cell death in individuals recovering from COVID-19. (A) The percentages of early apoptosis (Annexin V+ 7-AAD-) and late apoptosis / necroptosis (Annexin V+ 7-AAD+) of NKT-like cells in the blood of CI (n=30) compared to those of UI (n=21) were analyzed by flow cytometry. (B) Correlation analysis between the frequencies and the late apoptosis / necroptosis of NKT-like cells was performed in CI. (C) The percentages of early apoptosis (Annexin V+ 7-AAD-) and late apoptosis / necroptosis (Annexin V+ 7-AAD+) of CD4 T, CD8 T, B, and NK cells in the blood of CI (n=30) compared to those of UI (n=21) were analyzed by flow cytometry. CI: COVID-19 convalescent individuals, UI: SARS-CoV-2-unexposed individuals. Black and blue lines in the figures: median and quartiles. Statistically significant differences are indicated by asterisks (* < 0.05, ** <0.01; Non-parametric Mann-Whitney test).

Taken together, these results demonstrate that SARS-CoV-2 infections elicit a sustained impact on the immune cell composition in the peripheral blood during the extended convalescence phase, dominated by a contraction of NKT-like cells, and an expansion of DCs.

### Characterization of T cell phenotypes in individuals recovering from COVID-19

Next, we used several markers of CD4 and CD8 T cells to determine their differentiation (CD45RA and CCR7), proliferation (Ki67), activation (CD38 and HLA-DR), and exhaustion / suppression (PD-1, TIM-3, TOX, and Tregs) status. We also defined different subpopulations and T cells from UI and CI were divided into naïve (CD45RA+ CCR7+), central memory (TCM, CD45RA- CCR7+), effector memory (TEM, CD45RA- CCR7-), and terminally differentiated effector (TEMRA, CD45RA+ CCR7-) subpopulations (Figure S2). No significant differences were observed between UI and CI for any of the CD4 or CD8 T cell subpopulations mentioned above, although a tendency of decreased CD4+ TEMRA cell frequencies in CI (median: 3.3%) compared to UI (median: 7.5%) was observed (Figure S2). The proliferation (Ki67+) expression of both CD4 and CD8 T cells in CI was higher than that in UI (median: CD4 3.8% vs 2.7%, CD8 2.5% vs 1.9%), and the difference for CD8 T cells was statistically significant (Figure 3A), indicating that T cells from CI show an enhanced proliferation capacity. Previous studies have shown that T cells are highly activated during the acute phase of COVDI-19 (15), thus we next analyzed the activation status of CD4 and CD8 T cells by examining CD38 and HLA-DR expression on the cell surface. No significant differences in CD38 and HLA-DR expression on CD4 T cells were observed between UI and CI (Figure 3B and S3A). Compared to UI, CI showed 1.47-fold increase in the frequencies of CD38+ HLA-DR- CD8 T cells, however, this difference was not statistically significant (Figure 3C and S3A). Based on the analysis of PD-1 expression, some studies reported that CD8 T cells may already become functionally exhausted during the acute phase of COVID-19 (16), which was questioned by a different study from our group (Westmeier J, et al. In submission). In our current study, the PD-1 expression levels on CD4 or CD8 T cells from CI were similar to those in UI (Figure 3B, 3C and S3B). However, the expression of TIM-3, another immune check point molecule, was increased about 20% on CD4 and CD8 T cells in CI compared to UI, and this difference was statistically significant (Figure 3D). Moreover, we examined the expression of TOX in T cells from our study subjects, which is a newly identified key factor of T cell exhaustion (17, 18). The frequencies of TOX+ CD4 and CD8 T cells in CI increased around 20–30% compared to those in UI, however, the differences were not statistically significant (Figure 3B, 3C and S3B). Regulatory T cells (Tregs) play a very important role in controlling immunopathogenic reactions upon infections by dampening pathogen-specific immune responses (19–21). We therefore examined the frequencies of Tregs in the PBMCs of CI by analyzing Foxp3 expression in CD4 T cells. As shown in figure 3E, CI showed a significant increase in Tregs frequencies (median: 8.8%) compared to those in UI (median: 6.8%).

**Figure 3.**
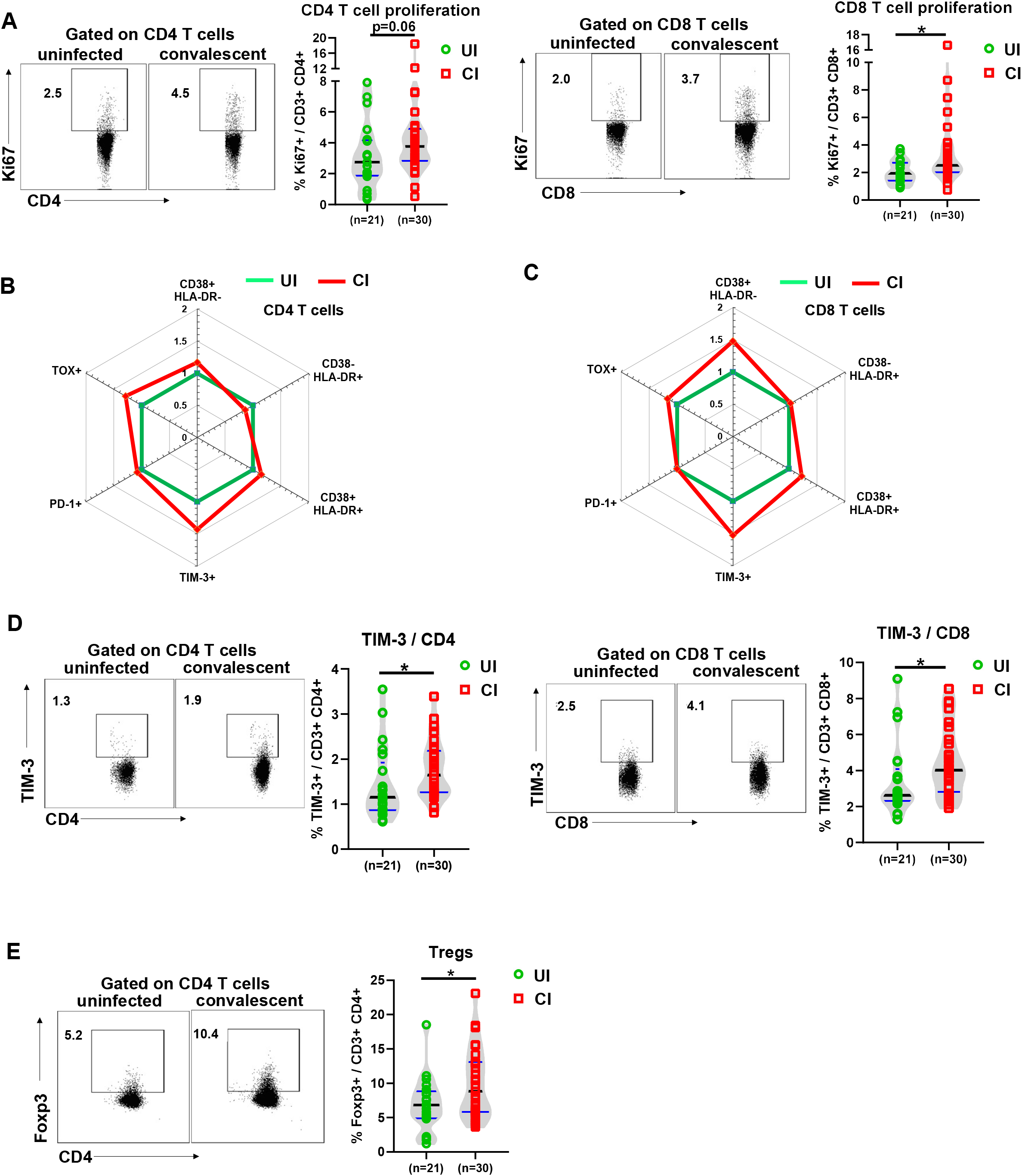
Characterization of T cell phenotypes in the PBMCs of individuals recovering from COVID-19. (A) The percentages of Ki67+ CD4 and CD8 T cells in the blood of UI (n=21) and CI (n=30) were analyzed by flow cytometry. (B) and (C) The fold changes of the percentages (median) of CD38+ HLA-DR-, CD38-HLA-DR+, CD38+ HLA-DR+, PD-1+ TOX-, PD-1- TOX+, PD-1+ TOX+, and TIM-3+ CD4 and CD8 T cells in the blood of CI (n=30) compared to those of UI (n=21) are depicted by radar plots. (D) The percentages of TIM-3+ CD4 and CD8 T cells in the blood of UI (n = 21) and CI (n=30) were analyzed by flow cytometry. (E) The percentages of Foxp3+ CD4 T cells in the blood of UI (n=21) and CI (n=30) were analyzed by flow cytometry. CI: COVID-19 convalescent individuals, UI: SARS-CoV-2-unexposed individuals. Black and blue lines in the figures: median and quartiles. Statistically significant differences are indicated by asterisks (* < 0.05, ** <0.01; Non-parametric Mann-Whitney test).

Taken together, our results demonstrate an immune environment that is prone towards T cell suppression during the late COVID-19 convalescent phase. However, T cells in CI still show a slightly enhanced activation and proliferation status, suggesting that these individuals are situated in a phase of ongoing restoration of the immune homeostasis.

### Characterization of cytotoxic effector profiles of T, NK, and NKT-like cells in individuals recovering from COVID-19

To characterize their cytotoxic profiles, we intracellularly stained CD4, CD8, NKT-like, and NK cells for the cytotoxic molecules Granzyme B (GzmB) and perforin directly *ex vivo* without re-stimulation and compared UI with CI. CI showed significant decreases in the frequencies of GzmB-producing NKT-like cells (median: 53.2%) and CD8 T cells (mean: 20.3%) compared to UI (NKT-like cells: 81.2%, CD8 T cells: 29.8%, figure 4A–4D). A tendency of decreased frequencies of GzmB-producing CD4 T cells was also observed, however, this difference was not statistically significant (Figure 4E and 4F). Consistently, the level (mean florescence intensity; MFI) of GzmB expression in individual NKT-like, CD4 and CD8 T cells was also significantly lower in CI than in UI (Figure 4A–4F). However, we did not observe significant differences in GzmB expression in NK cells. Interestingly, perforin expression was not different for all analyzed cell populations between CI and UI (Figure S4). We also examined the production of inflammatory cytokines such as interferon-γ IFN-γ), IL-6, and granulocyte-macrophage colony-stimulating factor (GM-CSF) by T cells, NK cells, and NKT-like cells in CI, since a previous study demonstrated that significant numbers of T cells produce these cytokines during the acute phase of COVID-19 (22). Although certain increases in the frequencies of CD4 and CD8 T cells producing IFN-γ, IL-6, and GM-CSF were observed in CI compared to UI (Figure 4Cand 4E), these differences were not statistically significant and appeared to be affected by two outliers in the CI group that had profound numbers of cytokine-producing T cells (Figure S4B and S4C). No significant differences in frequencies of IFN-γ, IL-6 and GM-CSF producing NK and NKT-like cells were observed between CI and UI (Figure S4A and S4F).

**Figure 4.**
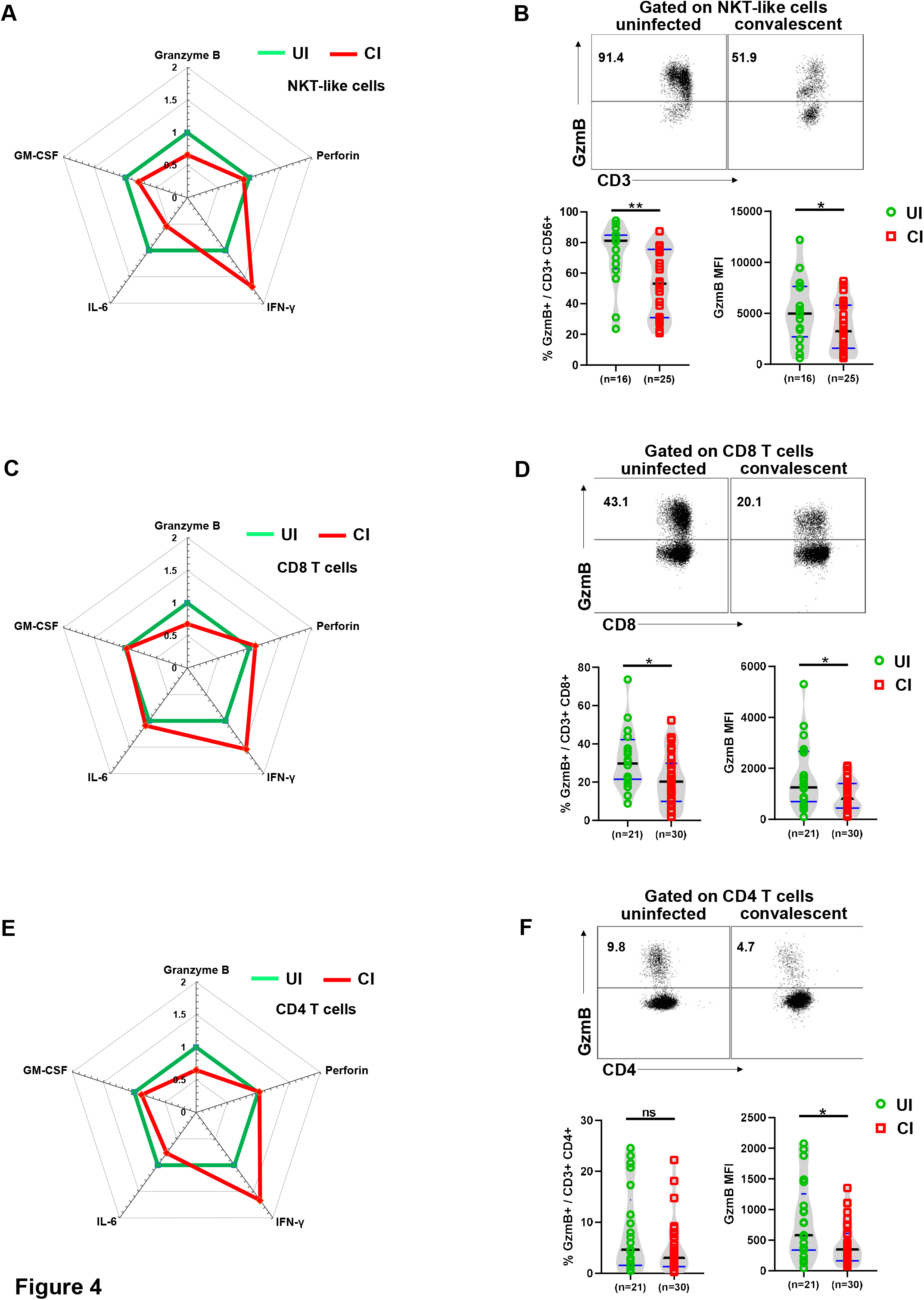
Characterization of cytotoxic and cytokine profiles of T, NK and NKT-like cells in the PBMCs of individuals recovering from COVID-19. (A) The fold changes of the percentages (median) of Granzyme B, perforin, IFN-γ, IL-6 and GM-CSF producing NKT-like cells in the blood of CI compared to those of UI are depicted by radar plots. (B) The percentages and MFI (Geometric mean) of Granzyme B expression of NKT-like cells in the blood of UI (n=16) and CI (n=25) were analyzed by flow cytometry. (C) The fold changes of the percentages (median) of Granzyme B, perforin, IFN-γ, IL-6 and GM-CSF producing CD8 T cells in the blood of CI compared to those of UI are depicted by radar plots. (D) The percentages and MFI (Geometric mean) of Granzyme B expression of CD8 T cells in the blood of UI (n=21) and CI (n=30) were analyzed by flow cytometry. (E) The fold changes of the percentages (median) of Granzyme B, perforin, IFN-γ, IL-6 and GM-CSF producing CD4 T cells in the blood of CI compared to those of UI are depicted by radar plots. (F) The percentages and MFI (Geometric mean) of Granzyme B expression of CD4 T cells in the blood of UI (n=21) and CI (n=30) were analyzed by flow cytometry. CI: COVID-19 convalescent individuals, UI: SARS-CoV-2-unexposed individuals. Black and blue lines in the figures: median and quartiles. Statistically significant differences are indicated by asterisks (* < 0.05, ** <0.01; Non-parametric Mann-Whitney test).

To further characterize effector functions of T cells in response to TCR stimulation. PBMCs from 5 CI and 5 UI were stimulated with anti-CD3/anti-CD28 for 5 days and were examined for cell proliferation (Ki67) and effector cytokine expression (IFN-γ. IL-2, and TNF-α). Compared to unstimulated cells, anti-CD3/anti-CD28 stimulation induced expected increases of Ki67 expression, as well as IFN-γ, IL-2, and TNF-αIL-2, and TNF-α production of CD4 and CD8 T cells in both CI and UI (Figure 5). No significant differences in effector cytokine production or proliferation of CD4 and CD8 T cells were observed between the groups (Figure 5B–5D).

**Figure 5.**
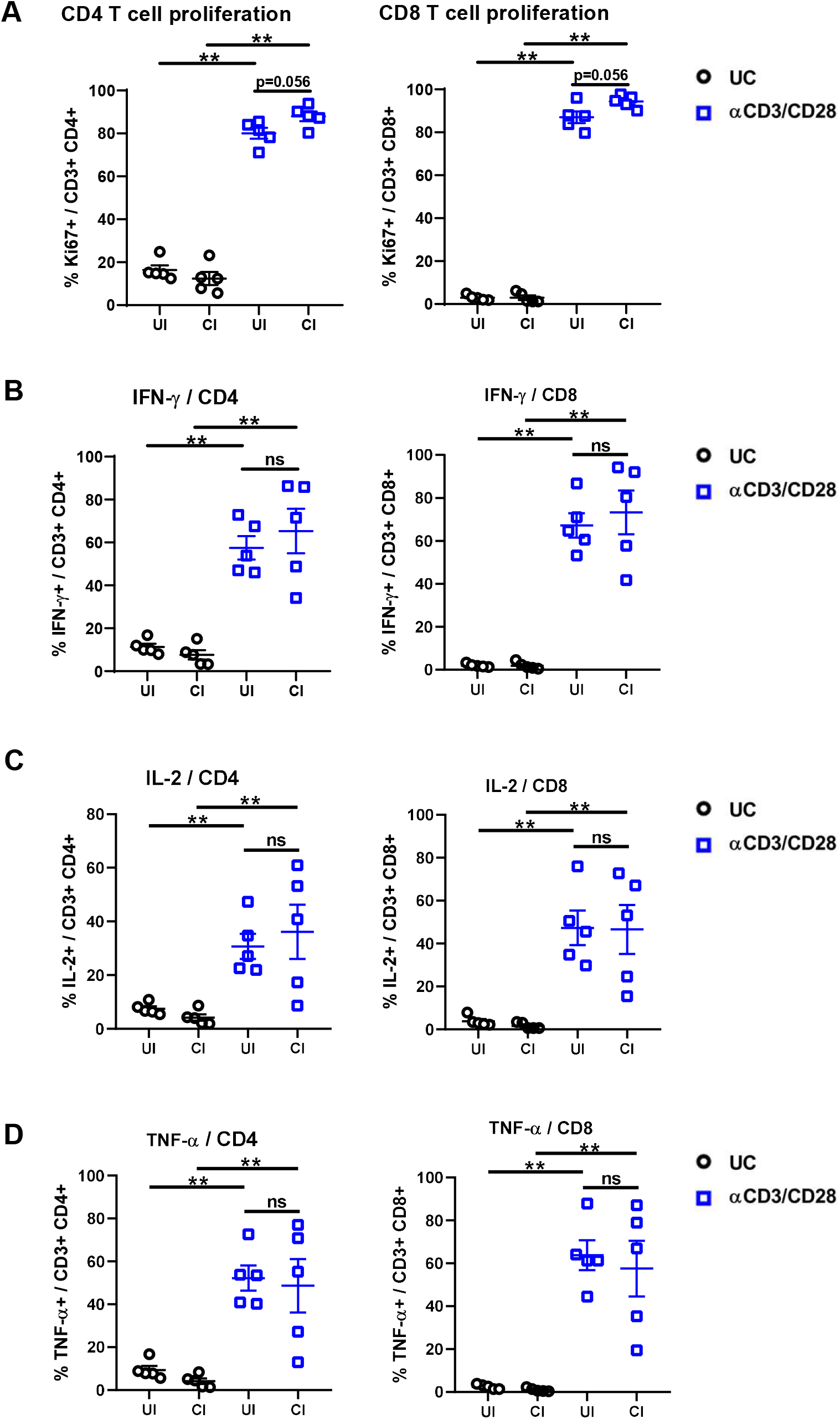
Characterization of the effector function of CD4 and CD8 T cells in the PBMCs of individuals recovering from COVID-19. PBMCs of UI (n=5) and CI (n=5) were either stimulated with anti-CD3 and anti-CD28 antibodies (α or left unstimulated (UC) and cultured for 5 days. The percentages Ki67 (A), IFN-α (B), IL-2 (C), and TNF-α (D) positive CD4 (left) and CD8 (right) T cells were analyzed by flow cytometry. CI: COVID-19 convalescent individuals, UI: SARS-CoV-2-unexposed individuals. Statistically significant differences are indicated by asterisks (* < 0.05, ** <0.01; Non-parametric Mann-Whitney test).

Taken together, these results indicate that there is a long-term suppression of the cytotoxic potential of T cells after resolving SARS-CoV-2 infection, however, general effector functions of T cells in COVID-19 convalescent individuals are maintained.

### Phenotype of antigen presenting cells in individuals recovering from COVID-19

Next, we evaluated the phenotype and functional properties of antigen presenting cells (APCs), including DCs, B cells, and monocytes by analyzing their CD80, CD86, CD72, and PD-L1 expression and compared UI with CI. DCs of CI showed about 20% increased CD80 expression intensities on the cell surface compared to those of UI and this difference was statistically significant (Figure 6A and 6B). CD80 and PD-L1 expression significantly increased by about 20% and CD72 by about 50% on the surface of B cells of CI (Figure 6C and 6D). No significant differences in CD86, CD72, and PD-L1 expression on DCS, CD86 on B cells, and CD80, CD86, CD72, and PD-L1 on monocytes were observed between the groups (Figure S5). Taken together, increased expression of costimulatory molecules on DCs and B cells suggests that APCs in CI are at a slightly enhanced activation status.

**Figure 6.**
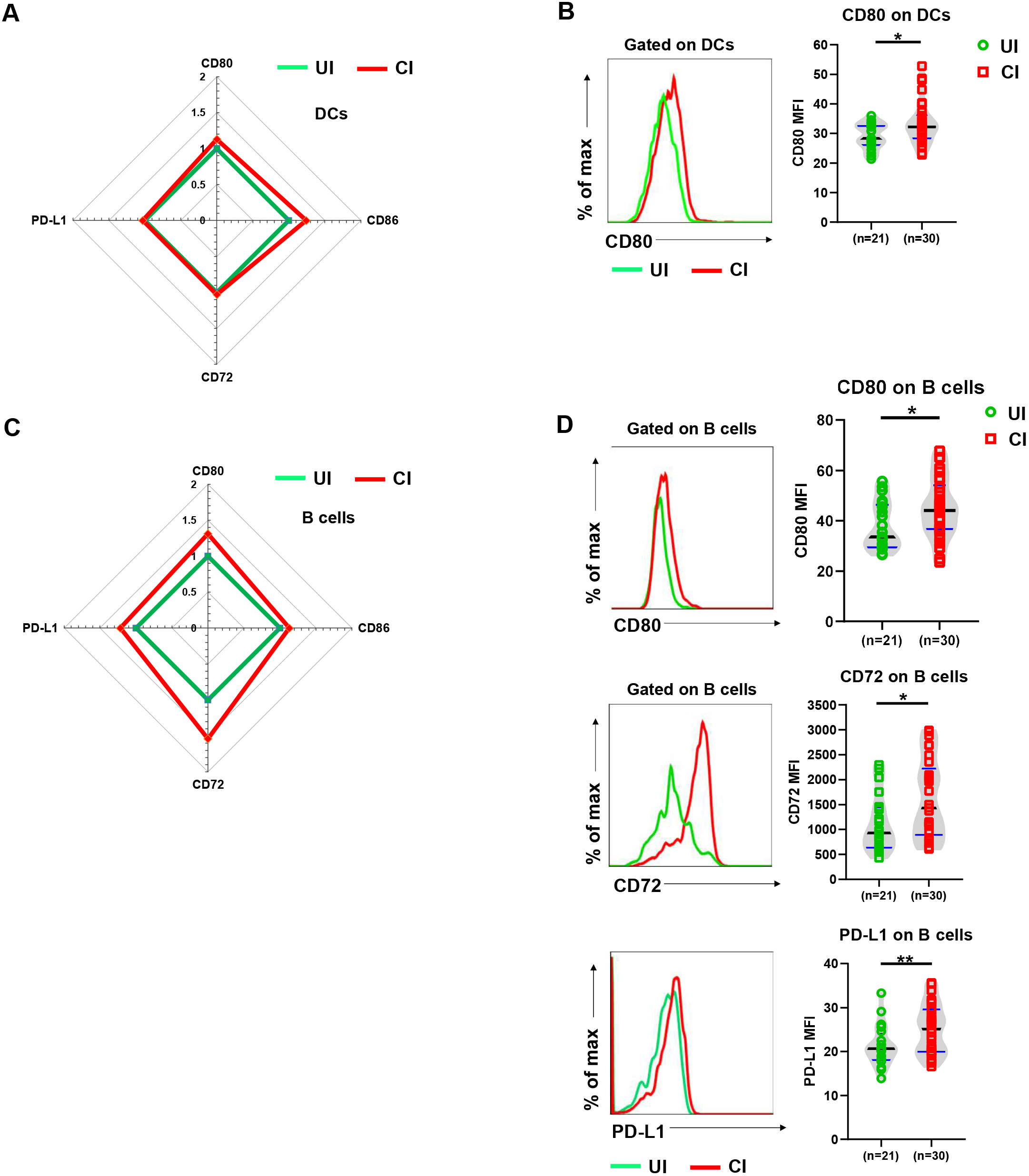
Characterization of phenotypes of antigen presenting cells in the PBMCs of individuals recovering from COVID-19. (A) The fold changes of the MFI (Geometric mean) of CD80, CD86, CD72, and PD-L1expression on DCs in the blood of CI (n = 30) compared to those of UI (n = 21) are depicted by radar plots. (B) The MFI (Geometric mean) of CD80 expression on DCs in the blood of UI (n = 21) and CI (n=30) were analyzed by flow cytometry. (C) The fold changes of the MFI (Geometric mean) of CD80, CD86, CD72, and PD-L1expression on B cells in the blood of CI (n=30) compared to those of UI (n = 21) are depicted by radar plots. (D) The MFI (Geometric mean) of CD80, CD72 and PD-L1 expression on DCs in the blood of UI (n=21) and CI (n=30) were analyzed by flow cytometry. CI: COVID-19 convalescent individuals, UI: SARS-CoV-2-unexposed individuals. Black and blue lines in the figures: median and quartiles. Statistically significant differences are indicated by asterisks (* < 0.05, ** < 0.01; Non-parametric Mann-Whitney test).

## Discussion

Wuhan was the very first city hit by SARS-CoV-2. Accordingly, the patients who experienced the longest phase of convalescence following COVID-19 reside here. This enabled us to investigate the ‘immunological scar’ left by SARS-CoV-2 on cellular immunity after recovery from the disease. Our results reveal that 2 to 4 months after resolved SARS-CoV-2 infection, most components of cellular immunity returned to normal. However, the previous SARS-CoV-2 infection could still be recognized during convalescent phase by diminished numbers of NKT-like cells and iNKT cells as well as increased DCs. CIs show an immune environment prone to suppression, supporting by the observation of significantly increases Treg frequencies, and upregulation of TIM-3 expression on T cells, and PD-L1 expression on B cells. Accordingly, the cytotoxic potential, as represented by GzmB expression, of T cells and NKT-like cells was significantly suppressed in CIs. Both CD4 and CD8 T cells of CIs showed increased cell proliferation and were fully capable of producing effector cytokines in response to TCR stimulation, suggesting the effector function of T cells is not compromised in CIs.

Unexpectedly, our study revealed profound changes of NKT cells in the convalescent phase of COVID-19. NKT cells are a small but important subset of T lymphocytes that regulate immune responses in the context of infection, cancer, and autoimmunity(23). NKT cells can promote cell-mediated immunity to tumors and pathogens, yet they can also suppress the cell-mediated immunity associated with autoimmune diseases and are involved in the pathogenesis of many inflammatory disorders (24).

NKT-like cells were shown to be cytotoxic toward lung epithelial cells and involved in the immunopathogenesis of pulmonary disease (25). It remains unclear by which means NKT cells carry out such opposing functions. The existence of functionally distinct NKT cell subsets may provide a rational explanation: so far, 3 NKT cell subsets, including classical NKT cells (iNKT cells), non-classical NKT cells, and NKT-like cells, each expressing different TCRs have been described (26). These cells are activated by lipid antigens linked to non-polymorphic CD1 molecules and/or pro-inflammatory cytokines generated during infection, and significantly contribute to the onset of infectious or autoimmune diseases (27). Reduced numbers of iNKT cells among PBMCs appear to correlate with the activity of systemic lupus erythematosus (SLE) disease (28). Selective loss of iNKT cells has also been reported during acute lymphocytic choriomeningitis virus (LCMV) infections (29), as well as in chronic HIV and UIV infection (30–32). Interestingly, subsequent long-term loss of iNKT cells during the convalescent phase following acute LCMV infection of mice has also been reported (33). It is believed that the reduction in iNKT cells at these late stages post-infection occurred by activation-induced cell death, since concomitant with the decrease in iNKT cells was an increase in the frequency of Annexin V+ iNKT cells(33). Highly similar to the observation in acute LCMV infections of mice, we also demonstrate a selective long-term loss of iNKT and NKT-like cells three months after recovering from an acute SARS-CoV-2 infection. The observation of increases in the frequency of Annexin V+ NKT-like cells in convalescent individuals suggests that the reduction of these cells may also occurred by activation-induced cell death. Recent studies have reported that during acute SARS-CoV-2 infection, NKT-like cells showed a significant increase in GzmB and perforin production (34), as well as a decrease in numbers in severe COVID-19 cases (35), suggesting these cells are highly activated during the acute phase. A very recent study has also demonstrated the expansion of NKT CD160 cluster in moderate but not severe COVID-19 patients, which was believed to promote rapid control of the disease through direct cytotoxicity as well as mediating the antibody-dependent cell-mediated cytotoxicity effect (36). Taken together, these data suggest that the activation and subsequent long-term loss of NKT and NKT-like cells during COVID-19 is a normal component of the host’s antiviral immune response. The mechanisms that regulate numbers and functions of NKT and NKT-like cells following SARS-CoV-2 infection should be further investigated.

In summary, we characterized the long-term impact of SARS-CoV-2 infection on the immune system and provide comprehensive picture of cellular immunity of a convalescent COVID-19 patient cohort with the longest recovery time. The overall alterations affecting cellular immunity observed in this study suggests that the immune system in convalescent individuals is going through a phase of restoring homeostasis after being highly activated during the acute phase of SARS-CoV-2 infection.

## Data Availability

The availability of all data referred to in the manuscript must obtain permission from the corresponding author.

## Acknowledgement

This work is supported by the Fundamental Research Funds for the Central Universities (2020kfyXGYJ028, 2020kfyXGYJ046 and 2020kfyXGYJ016), the National Natural Science Foundation of China (81861138044 and 91742114), the National Science and Technology Major Project (2017ZX10202203), and the Medical Faculty of the University of Duisburg-Essen and Stiftung Universitaetsmedizin, University Hospital Essen, Germany.

